# Impaired everyday executive functions and cognitive strategy use on the Weekly Calendar Planning Activity in individuals with stroke undergoing acute inpatient rehabilitation

**DOI:** 10.1101/2021.05.03.21256540

**Authors:** Abhishek Jaywant, Catherine Arora, Alexis Lussier, Joan Toglia

## Abstract

Executive dysfunction is common in stroke patients undergoing acute inpatient rehabilitation. However, comprehensive neuropsychological evaluation of executive functions is often not feasible in this setting. Objective, standardized, and performance-based measures of cognitively-based instrumental activities of daily living (C-IADL) can capture everyday executive functions and inform rehabilitation planning and interventions. The goal of this study was to compare performance of individuals with stroke to healthy age-matched adults in performance on the 10-item Weekly Calendar Planning Activity (WCPA). 77 stroke inpatients undergoing rehabilitation and 77 healthy control participants completed the WCPA, a C-IADL measure involving scheduling appointments that requires planning, working memory, shifting, and inhibitory control. Compared to the control group, stroke patients had significantly worse accuracy, made more errors, used fewer cognitive strategies, followed fewer rules, took more time to complete the task, and were less efficient. 83% of stroke patients were less accurate than predicted by their age, and 64% used less strategies than their age prediction. Among 28 participants who screened as having “normal” cognitive function on the Montreal Cognitive Assessment, the majority had deficits on the WCPA. Our results indicate that the WCPA is sensitive to executive dysfunction in stroke patients undergoing acute inpatient rehabilitation and underscores deficits in C-IADL accuracy, efficiency, and strategy use at this stage of stroke recovery. The WCPA may be a useful measure to ascertain executive dysfunction and to incorporate in cognitive rehabilitation.

## Introduction

Cognitive impairments are common following stroke and contribute to limitations in daily activities and poor functional outcomes. Cognitive deficits are observed in approximately half of stroke survivors in the acute period (<1 month) post-stroke and persist long-term (1). Impairments in executive functions are especially common in the acute inpatient rehabilitation setting and associated with worse functional gain and greater limitations in activities of daily living after discharge (2–4).

Although neuropsychological testing remains the gold standard for assessing cognition at the impairment level, comprehensive neuropsychological evaluation is not always feasible in the acute inpatient rehabilitation setting. Functional cognitive assessments that objectively assess performance in cognitively-based instrumental activities of daily (C-IADL)— such as organizing a schedule, paying bills, or managing medications—can serve as a valuable complement to impairment-based evaluation of cognition. Performance-based and functionally relevant C-IADL measures typically have strong ecological validity and provide the patient and the examiner with perspective into the patient’s executive skills in a functional context, which are especially relevant in rehabilitation and to independent functioning (5). C-IADL measures can be particularly valuable in the acute inpatient rehabilitation setting for developing tailored cognitive rehabilitation interventions (6,7). C-IADL measures are known to add sensitivity to detecting executive dysfunction that is not captured by impairment-based cognitive tests (8,9).

The Weekly Calendar Activity (WCPA) (10,11) is an objective, standardized, and validated performance-based measure of C-IADL that assesses multiple aspects of a person’s executive skills in a functional, ecologically-valid context. The WCPA requires working memory, planning, shifting, inhibition, and self-monitoring to complete. It also assesses a person’s cognitive strategy use in facilitating efficient performance. The standard 17-item version of the WCPA differentiates between healthy controls and a wide range of populations with executive dysfunction including those with multiple sclerosis (12), mild cognitive impairment (13), attention-deficit hyperactivity disorder (14), pediatric acquired brain injury (15), and epilepsy (16). It is responsive to change in executive functions in individuals with schizophrenia undergoing a metacognitive cognitive rehabilitation program (17). Whether the WCPA can differentiate between healthy adults and individuals with stroke in evaluating executive dysfunction after stroke, and specifically in the inpatient rehabilitation setting with the shorter 10-item version, has to date not been established. Given that the WCPA relies on planning, working memory shifting, and inhibition—abilities that are frequently impaired post-stroke (18,19)— the WCPA may be sensitive to C-IADL deficits and differentiate patients from age-matched healthy adults in the acute inpatient rehabilitation setting.

The goal of this study was to compare individuals with stroke to healthy age-matched adults in performance on the 10-item short-form/inpatient version of the WCPA. We hypothesized that relative to the healthy control group, individuals with stroke would have lower percentage accuracy of appointments entered, and a lower number of strategies used, which are the primary outcomes of executive function/C-IADL and cognitive strategy use respectively on the WCPA. We also hypothesized that compared to healthy participants, stroke patients would spend less time planning, take longer to complete the task, follow fewer rules correctly, and use fewer cognitive strategies. We predicted that WCPA performance would be correlated only modestly with an impairment-level measure of cognition, given that there is only partial overlap between impairment-based and C-IADL measures of cognition (8). Finally, we explored whether the WCPA would be sensitive to executive dysfunction in individuals who screened as having normal cognitive functioning.

## Methods and Materials

### Participants

N=77 individuals with stroke and N=77 healthy age matched controls from a larger existing normative database were included in this study. Stroke patients were all undergoing acute inpatient rehabilitation on a 22-bed general rehabilitation unit at a large, urban academic medical center. Inclusion criteria were the same as for admission to the inpatient rehabilitation unit: medically stable for rehabilitation, ability to tolerate three hours of rehabilitation therapy daily, and reasonable expectation for functional gain. The 10-item WCPA was administered as part of standard of care on the inpatient rehabilitation unit by Occupational Therapists for persons who were alert, oriented, able to attend for at least 20 minutes, able to read and write legibly in English, follow two-step commands, and were cognitively independent in basic self-care activities of daily living (ADL). Exclusion criteria included those who would not be typically given the WCPA during ordinary care such as those with dementia, severe cognitive impairment, language or visual deficits, or required cognitive assistance for basic self-care activities. People with limited English proficiency were also excluded as the test materials were only available in English. All study procedures were approved by the Weill Cornell Medicine Institutional Review Board.

Healthy control participants were obtained from an existing normative database. Participants were recruited via snowballing techniques by graduate occupational therapy students from the greater New York City area. Inclusion criteria were those who were living independently in the community, and for participants age over 65, a score >24 on the Montreal Cognitive Assessment. Exclusion criteria were reported past history of a neurological condition, attention-deficit hyperactivity disorder, history of hospitalization for a psychiatric disorder, or inability to read or write in English. Collection of normative data from healthy controls was granted exemption by the Mercy College Institutional Review Board (IRB), because data were recorded such that participants could not be identified. An oral consent script was read aloud, and a written copy of the script was provided to each participant.

### Measures

#### Weekly Calendar Planning Activity

The WCPA is an objective, performance-based C-IADL measure of working memory, planning, shifting, inhibition, and self-monitoring. It has demonstrated validity, reliability, and sensitivity to executive dysfunction and sensitivity to change (7,10,11,20). The WCPA requires the examinee to input a series of appointments into a mock weekly calendar/schedule while following a set of specific rules and guidelines (Figure 1). Appointments are either fixed at a certain date/time or flexible, and at times conflict, which requires the examinee to plan an efficient approach. The examinee has to keep track of multiple rules (e.g., cannot enter appointments on a certain day, cannot cross off items once entered) in working memory while shifting between the appointment sheet, calendar, and instructions sheets. The examiner periodically attempts to distract the examinee with pre-specified questions, which the examinee has to inhibit. The examiner observes the examinee and records specific strategies that he or she uses; the examinee also reports to the examiner at the end of the task any additional strategies that he or she employed in a post-task interview.

**Figure 1.**
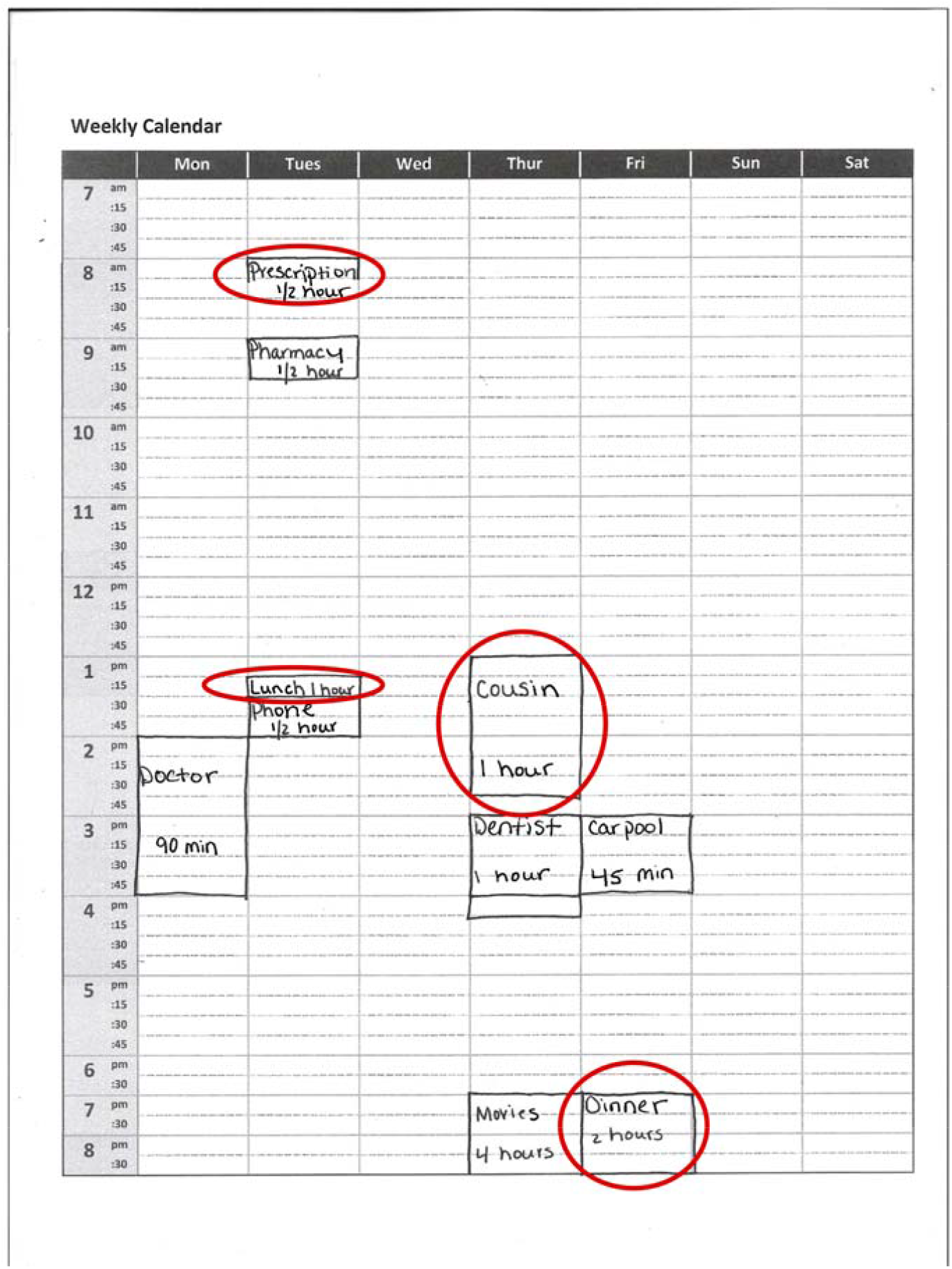
Visual example of the weekly calendar stimulus on the Weekly Calendar Planning Activity. Patients are required to schedule appointments of specific lengths on specific days and times while following multiple rules. Red circles highlight errors, which can include placing the appointment on the wrong day or time; marking the appointment with an incorrect duration; or having a vague description of the appointment.

In this study, we used the 10-appointment version of the WCPA. The WCPA-10 has the same ratio of fixed and variable appointments (3/7 or 70%) as the WCPA-17, but there are only 10 appointments to enter as opposed to 17. The main outcome measure was the percent of appointments entered correctly out of 10 (Percent Accuracy). We also calculated Planning Time (time in seconds from the start of the task to entering the first appointment), Time to Completion, Efficiency Score (time in seconds / weighted accuracy), Total Errors, the number of Rules Followed correctly out of 5, and Total Strategies (combination of those observed by the examiner and self-reported by the examinee). A lower efficiency score indicates that the client obtained higher accuracy in less time. Efficiency scores were not calculated for those with accuracy scores of 3 or below. Based on the standard WCPA record form, we also documented for each participant whether or not they used one of several different cognitive strategies.

#### Montreal Cognitive Assessment (MoCA)

The MoCA (21) is a 30-item screening measure for general cognitive impairment that is administered on admission as standard of care to all stroke patients on our acute inpatient rehabilitation unit. The MoCA assesses visuospatial/executive skills, naming, attention, language, abstraction, delayed recall, and orientation. Lower scores indicate greater cognitive impairment. The MoCA has demonstrated validity and clinical utility in inpatient stroke rehabilitation (22), and is closely associated with impairments assessed using neuropsychological tests (2).

### Statistical Analysis

We used one way analysis of variance (ANOVA) and chi-square tests to evaluate group differences in demographic and clinical variables. We used one-way ANOVAs to evaluate group differences on each of the outcome measures, Percent Accuracy, Planning Time, Time to Completion, Efficiency Score, Total Strategies, Total Errors, Rules Followed, and Total Strategies. For cognitive strategies that were commonly used by the healthy control group (at least n=20 [25%] of the control group used), we compared the frequency of use by individuals with stroke to healthy control participants using chi-square tests.

We next sought to evaluate individual differences in the performance of stroke participants relative to the healthy control group, correcting for demographic factors. We first used Spearman rank-order correlations to evaluate in the healthy control group the association between age, education, and gender and Percent Accuracy (as a measure of overall executive skills) and Total Strategies (as a measure of cognitive strategy use). We then used demographic-corrected regression equations—including predictors that exhibited significant correlations with Percent Accuracy and Total Strategies— to obtain the demographic-predicted score for each stroke participant. We subsequently subtracted each participant’s demographic-predicted score from his or her obtained score to obtain the residual demographic-corrected score. We reported the frequencies of these residual scores for the entire sample, and for those patients who scored in the normal range on the MoCA (25 or greater /30), the latter in order to explore the clinical utility of the WCPA in individuals with stroke who screen as having normal cognitive functioning.

## Results

### Demographics and clinical characteristics

There were no group differences in age, gender, or education (Table 1). There was a significant difference in race/ethnicity between groups. Both groups had similar percentages of Caucasian participants, while a greater percentage of Black participants and a smaller percentage of Hispanic participants were observed in the stroke group. Stroke participants had significantly lower MoCA scores than the healthy control group.

**Table 1.**
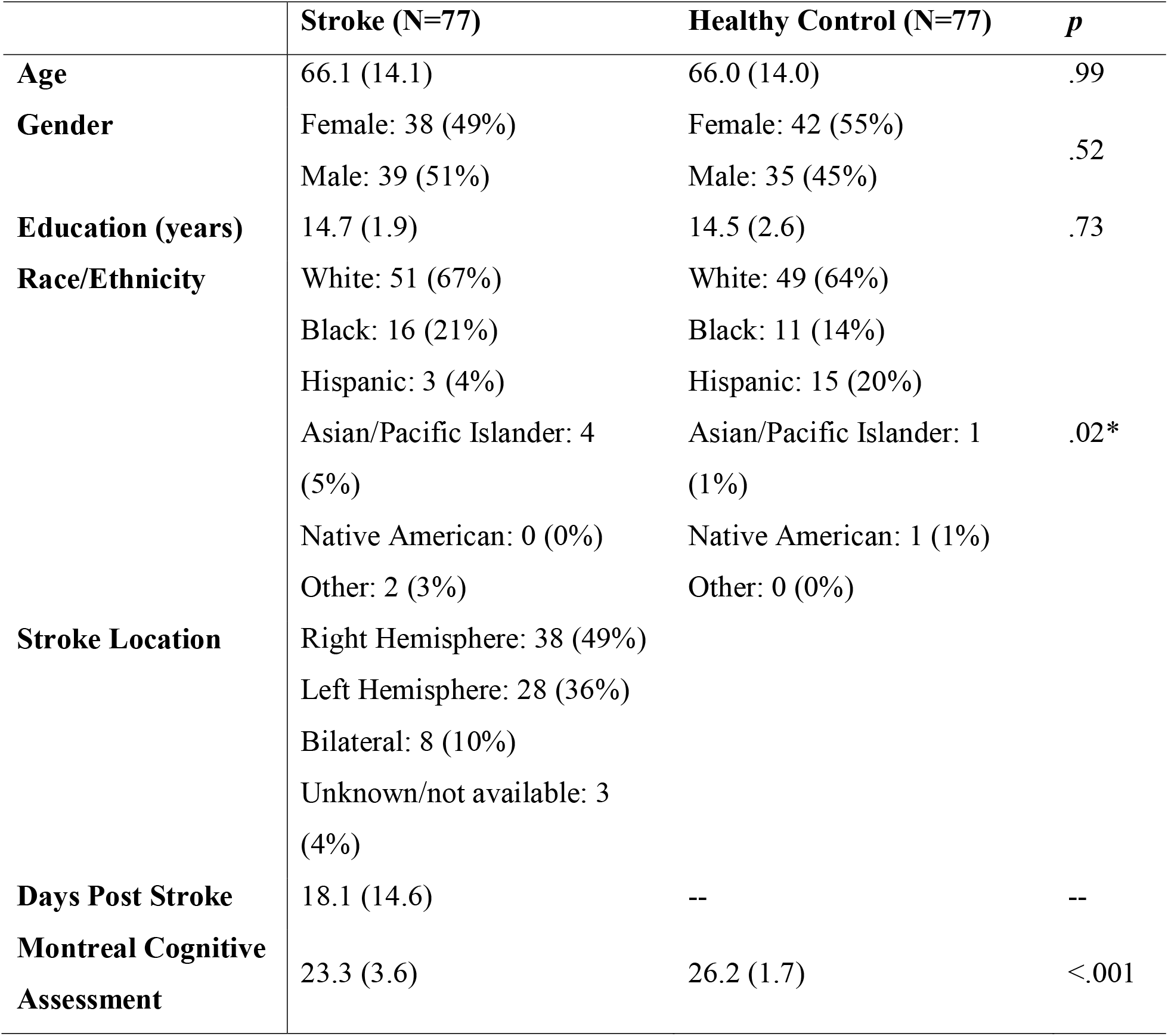
Demographic and clinical characteristics.

### Performance on the WCPA

On average, the WCPA took approximately 12 minutes for stroke participants to complete. Using one-way ANOVAs, relative to control participants, stroke patients had significantly worse Percent Accuracy, Total Strategies, Time to Completion, Efficiency Score, Rules Followed, and Total Errors (Table 2). Stroke patients and control participants did not differ in WCPA Planning Time.

**Table 2.** Performance on the WCPA by Group.

| **WCPA Measure** | **Stroke** | **Healthy Control** | **p-value** |
| --- | --- | --- | --- |
| **Percent Accuracy** | 49.9 (24.1) | 71.0 (18.3) | <.001 |
| **Total Strategies** | 3.9 (2.0) | 5.0 (2.5) | <.002 |
| **Planning Time (s)** | 89.5 (199.1) | 63.1 (75.4) | .29 |
| **Time to Completion (s)** | 767.1 (399.6) | 553.9 (196.0) | <.001 |
| **\*Efficiency Score** | 266.7 (242.3) | 120.6 (71.7) | <.001 |
| **Rules Followed** | 3.7 (1.0) | 4.2 (0.8) | <.001 |
| **Total Errors** | 5.0 (2.4) | 2.9 (1.9) | <.001 |

\*Higher scores indicate lower efficiency

The number of strategies used was significantly related to the percentage of accurate appointments on the WCPA (r=.37, p <.001). The following strategies were used by at least n=20 (25%) of the healthy control group: repeats keywords or instructions out loud; uses finger; crosses off, checks off, or highlights appointments entered; enters fixed appointments first and then flexible appointments; self-checks; talks out loud about strategy or plan; and pauses and rereads. Individuals with stroke less frequently used their finger, crossed/checked/highlighted appointments, entered fixed appointments first and then flexible appointments, and self-checked (Figure 2; all *X*^2^ > 6.9, p’s <.03). There was no group difference in frequency of repeating keywords/instructions out loud, or in frequency of pausing and rereading.

**Figure 2.**
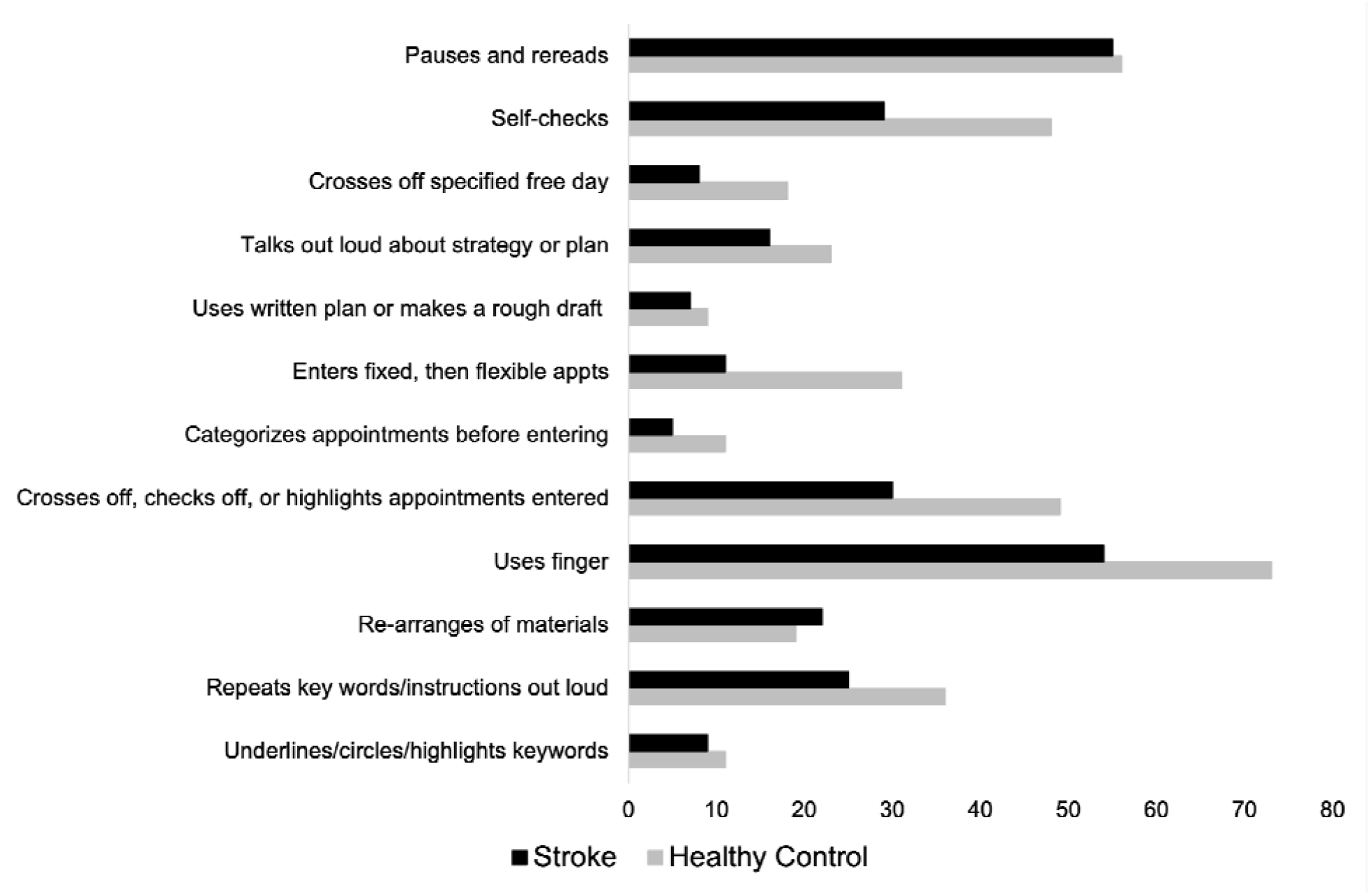
Frequency of strategies used by stroke patients and healthy control participants.

### Correlation with Cognitive Impairment

Performance on the MoCA was modestly but significantly correlated with Percent Accuracy (r_s_=.36, p<.001), Rules Followed (r_s_=.26, p=.004), and Total Strategies (r_s_=.31, p=.001), and Efficiency Score (r_s_=-.29, p=.004). MoCA score was not correlated with Time to Completion (r_s_=-.15, p=.11) or Planning Time (r_s_=-.04, p=.71).

### Individual Differences in Performance in Stroke Participants Relative to Control Group after Demographic Correction

In the healthy control group, Percent Accuracy correlated significantly with age (r_s_ = -.39, p<.001) but not education (r_s_ = .15, p=.22). Similarly, Total Strategies correlated significantly with age (r_s_ = -.52, p <.001) but not education (r_s_ = .19, p = .12). We thus computed regression equations predicting Percent Accuracy and Total Strategies from age. The relationship between Percent Accuracy and age was modeled by *y = 106*.*67 + (-*.*54)*(age)*, and the relationship between Total Strategies and age was modeled by *y = 10*.*52 + (-*.*08)*(age)*. Using these equations, we calculated each stroke participant’s age-predicted Percent Accuracy score and Total Strategies score, and subtracted these values from their obtained scores.

Results are displayed as box plots (median and interquartile range) in Figure 3, with negative values indicating performance worse than would be expected by age. As a group, stroke participants had a median Percent Accuracy 19.1% lower than would be predicted by age (range = 79.4% lower to 27.9% higher). 64/77 (83.1%) stroke participants were less accurate on the WCPA than their age prediction. Similarly, stroke participants as a group had a median Total Strategies 1.6 lower than would be predicted by age (range = 7 lower to 6 higher). 55/74 (74%) stroke participants used fewer strategies than their age prediction; three stroke participants were missing data on strategy use.

**Figure 3.**
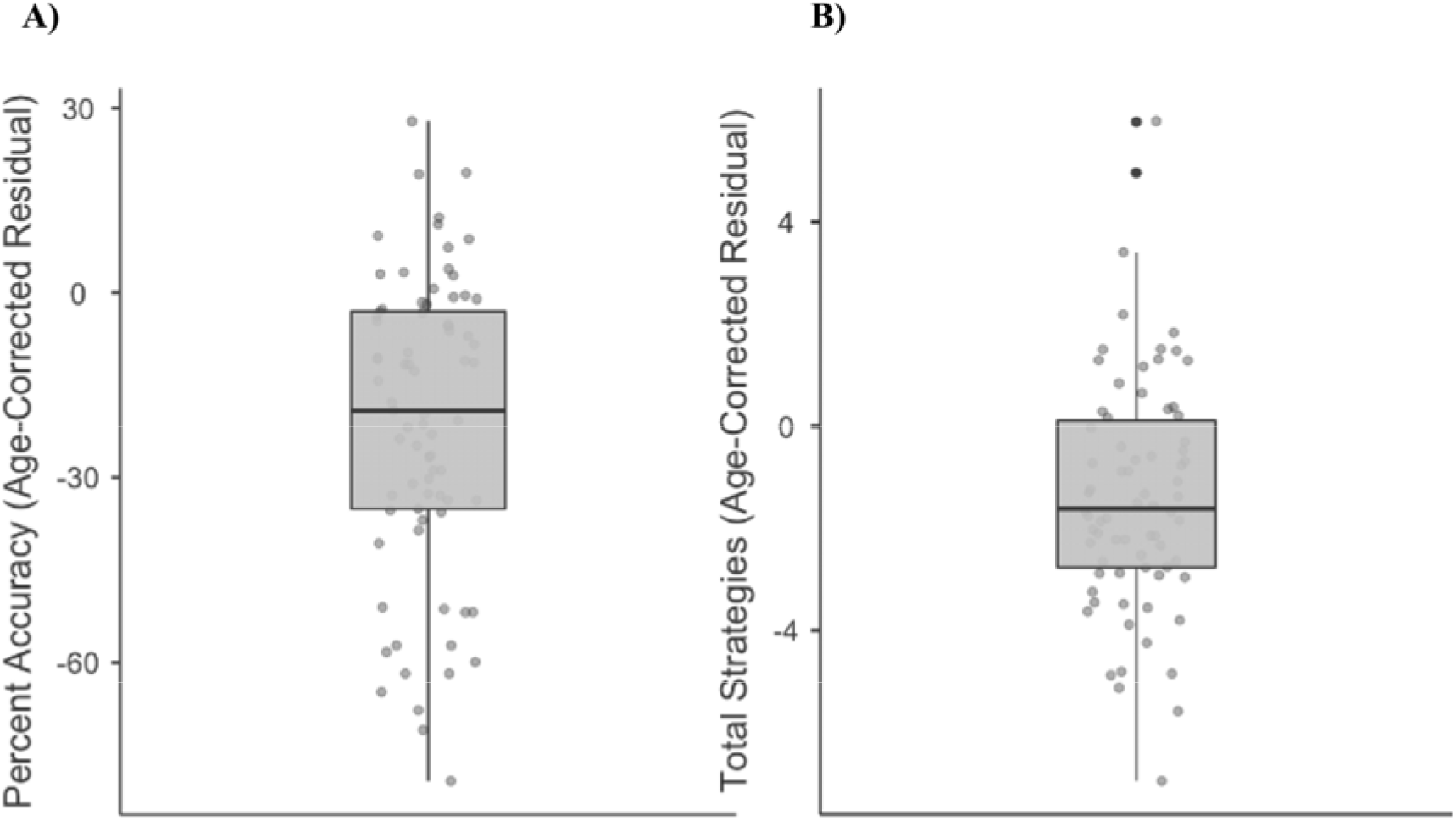
Boxplots showing median, interquartile range, range, and individual datapoints of stroke patient residual scores (raw score – age-predicted score) for percent accuracy (A) and total strategies (B). Median/interquartile range of residual scores are below age predictions.

We then explored individual differences in performance (Percent Accuracy and Total Strategies) using the regression-predicted and age-corrected procedure above, but in stroke participants who scored within normal limits (25/30 or higher) on the MoCA (Figure 4). Such participants would be classified clinically as having “normal” cognitive functioning based on standard of care cognitive screening on our inpatient rehabilitation unit. 28 individuals in our sample scored within normal limits on the MoCA. Within this subgroup, median Percent Accuracy was 11.2% lower than age prediction (range: 61.8% lower to 27.9% higher). 23/28 stroke participants (82.1%) performed below their age-predicted score in Percent Accuracy. Within this subgroup, median Total Strategies was 1.32 lower than predicted by age (range: 4.8 lower to 6 higher). 20/27 stroke participants (74.1%; 1 individual with missing data) performed below their age-predicted score in Total Strategies.

**Figure 4.**
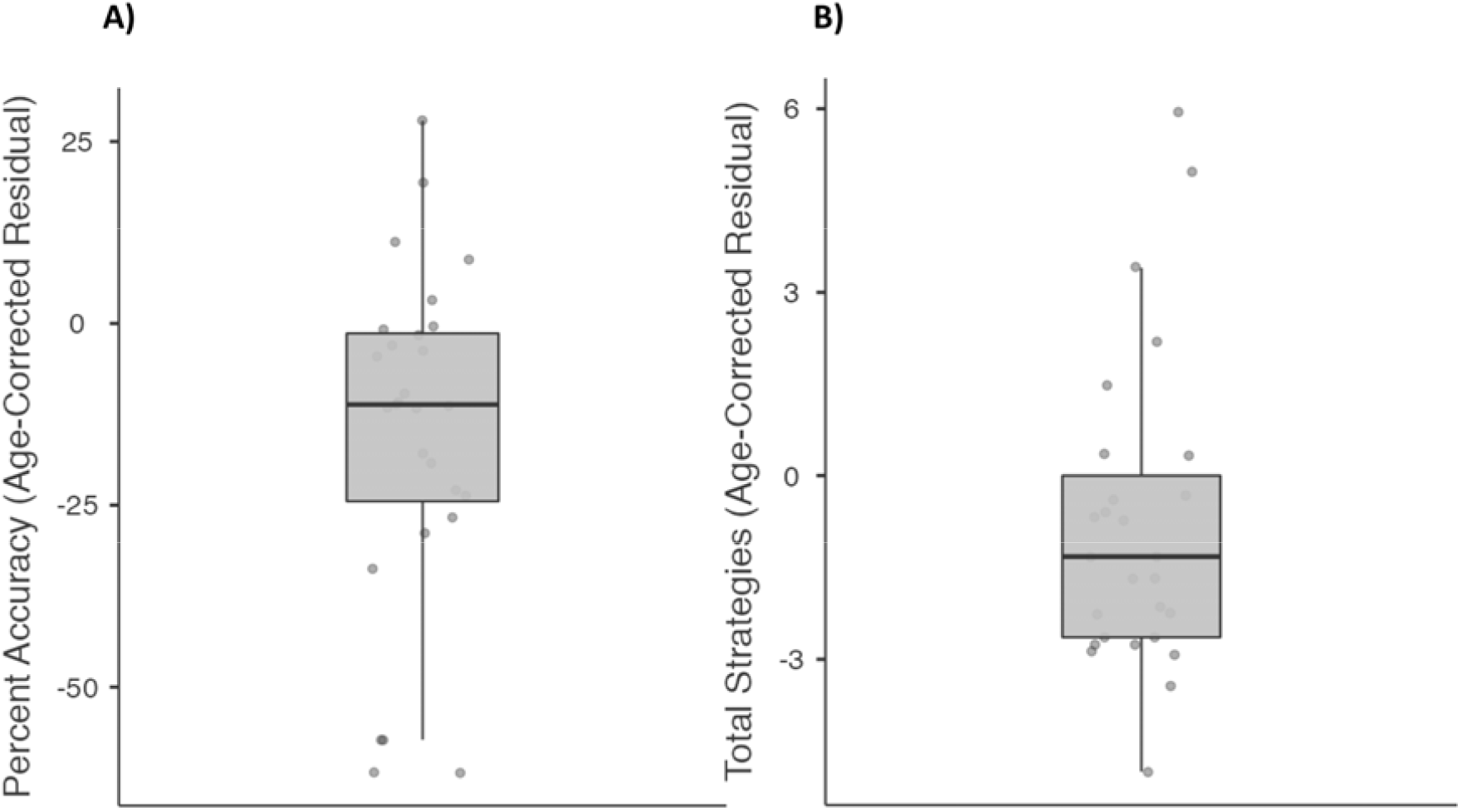
Boxplots showing median, interquartile range, range, and individual datapoints of stroke patient residual scores (raw score – age-predicted score) for percent accuracy (A) and total strategies (B), in patients deemed to have “normal” cognitive function on the Montreal Cognitive Assessment. Median/interquartile range of residual scores are below age predictions.

## Discussion

The main findings of this study are that (1) a standardized, objective, and performance-based measure of C-IADL and everyday executive functions, the WCPA, is feasible, quick to administer, and sensitive to executive dysfunction in individuals with stroke undergoing inpatient rehabilitation; (2) the WCPA is also sensitive to identifying poor cognitive strategy use by stroke patients relative to healthy control participants; (3) at an individual level, the majority of stroke patients score below their age-predicted performance on the WCPA, including overall accuracy and total strategies used; and (4) performance on the WCPA correlates only modestly with an impairment-based screening measure of cognition (MoCA) and identifies deficits in patients who would be deemed to have “normal” cognition based on the MoCA.

The WCPA differentiated individuals with stroke from healthy control participants on multiple aspects of performance and identified performance deficits that can be easily missed within a structured inpatient rehabilitation setting. Specifically, relative to the control group, our sample of stroke patients had significantly lower accuracy, followed fewer rules, made a greater number of errors, were less efficient, and took longer to complete the WCPA. Findings suggest that the WCPA is sensitive to stroke-related deficits in working memory, processing speed, error monitoring and resolution, and shifting/inhibition. These performance deficits are consistent with the known executive dysfunction that predominates post-stroke (REF), including in the acute period while patients are undergoing inpatient rehabilitation (REF). Interestingly, the stroke and control groups did not differ in planning time on the WCPA. That is, stroke patients on average did not take more or less time relative to control participants to plan their approach to the task, prior to initiating the first appointment entered. This may be because the WCPA goal of entering a list of appointments into a calendar appears deceptively easier than it actually is. Healthy control participants also demonstrated relatively brief planning times; however, they were observed to more frequently stop, pause and readjust task methods once they encountered potential appointment conflicts or recognized task complexities. Pause and stop periods within the task, may thus be better indications of planning than the initial planning time in this particular task.

An advantage of the WCPA is that it assesses both *number* and *type* of strategies used, enabling the objective quantification of cognitive strategy use. This is especially relevant in the inpatient rehabilitation setting where rehabilitation clinicians are teaching patients strategies to optimize performance and maximize independence in C-IADLs in preparation for discharge back to the community. We found that individuals with stroke less frequently used particular types of cognitive strategies on the WCPA. Specifically, they less frequently used their finger (i.e., to focus and maintain attention on salient aspects of the stimuli), less frequently crossed out/checked off/highlighted appointments to keep track of those that had been entered and those that had not been entered, less often entered fixed appointments first and then flexible appointments, and less frequently self-checked for errors. The lower use of these external strategies likely increased demands on working memory and cognitive load, thereby contributing to worse performance. This supports literature suggesting that strategy training may be an important ingredient in enhancing functional cognitive abilities following a stroke or acquired brain injury (7,23). Further, limited use of self-checking strategies implies that some participants may not have been aware of performance challenges or the need to evaluate, monitor, and adjust task methods. Careful analysis of strategy deficiencies within the context of a functional cognitive task can inform the types of strategies and training that may be most useful for clinicians to emphasize during rehabilitation. This finding also highlights the benefit of a measure such as the WCPA, which is both sensitive to identifying functional cognitive deficits and provides clinicians with readily usable information to guide rehabilitation treatment and discharge planning.

Another goal of this study was to investigate, at an individual level, how individuals with stroke performed relative to demographic expectations. Given significant associations between age and WCPA performance, we derived age-predicted scores for each stroke participant and subtracted their raw score from their predicted score. We found that the majority of stroke patients (83%) had performed worse on the WCPA than their age prediction. Further, the majority of stroke patients (74%) used fewer cognitive strategies than their age prediction. This analysis highlights deficiencies in everyday executive functions and cognitive strategy use in stroke patients at the individual level, in addition to at the group level. Findings also highlight a large inter-individual range of scores relative to age predictions (as shown in Figure 3), suggesting that some patients perform well below age expectations while others may surpass age expectations. Future studies should evaluate predictors and correlates of individual WCPA performance. Clinicians may find it useful to incorporate our regression equations to compare individual patient performance to that patient’s age-predicted score.

We found only modest correlations between the WCPA and the MoCA. This finding accords with research indicating only partial overlap between impairment-based and functional measures of cognition (8,24). It highlights the complementary value of measures such as the WCPA that focus on C-IADLs and everyday executive functions in an ecologically-valid manner.

Importantly, the WCPA identified executive functioning deficits and worse cognitive strategy use in patients who scored within the normal range on the MoCA. 82% of patients classified as “normal” on the MoCA had worse accuracy than their age prediction and 74% used fewer strategies than their age prediction. This finding further underscores the utility of the WCPA as an adjunct to impairment-based cognitive screening measures such as the MoCA.

### Limitations

Our characterization of clinical stroke characteristics was relatively limited. Because our data were collected in the context of routine clinical care, this limited the ability to collect more comprehensive information such as stroke location or type, stroke severity, or medical comorbidities. However, this reflects the realities of clinical research in an acute inpatient rehabilitation setting. Future work on the WCPA will benefit from investigating the relationship between clinical-disease characteristics and performance. Relatedly, the MoCA is a relatively brief screening measure of cognitive impairment. Our stroke sample was not routinely administered comprehensive neuropsychological measures of executive functioning and other cognitive domains to which we could compare performance on the WCPA. However, this reflects the reality of integrating assessments on acute inpatient rehabilitation units in which it is not always feasible to conduct extensive neuropsychological testing.

### Conclusion

The WCPA, a multi-step functional cognitive (C-IADL) task is feasible in an inpatient setting, quick to administer, and appears sensitive to subtle executive dysfunction, even for those who perform above the normal cut-off score on a cognitive screening tool. Relative to healthy adults, individuals with stroke, perform significantly worse on the WCPA and use significantly fewer cognitive strategies, both at the group level and commonly on an individual level. This finding emphasizes the importance of analyzing deficiencies in cognitive strategy use and considering methods for promoting strategy use within rehabilitation.

C-IADL skills are typically under-assessed in inpatient rehabilitation settings in people with stroke due to time constraints and a focus on physical abilities and self-care skills. This paper is the first to report findings of the 10-item version of the WCPA, thereby contributing to the limited (scarce) literature on C-IADL assessment and strategy use in those with mild cognitive deficits and stroke. The results highlight the need to routinely use a higher level functional cognitive assessment tool like the WCPA, for those that are cognitively independent in self-care ADL in an inpatient setting, to identify difficulties that may interfere with safety and independence upon discharge to the home and community.

## Data Availability

Data for this manuscript are not available.

## Acknowledgements

The authors received no specific funding for this project. AJ receives salary support from a K12 career development award from Georgetown University/National Institute of Child Health and Human Development (Grant/Award Number: 1K12□HD093427□04). The authors declare that the research was conducted in the absence of any commercial or financial relationships that could be construed as a potential conflict of interest. A portion of these results was presented at the 2019 annual meeting of the American Occupational Therapy Association. We thank Michael W. O’Dell, M.D., for his support of this research and Gargi Doulatani, M.A. for help with data extraction.

## References

1. Turunen KEA, Laari SPK, Kauranen TV, Uimonen J, Mustanoja S, Tatlisumak T, et al. Domain-specific cognitive recovery after first-ever stroke: A 2-year follow-up. J Int Neuropsychol Soc. 2018;24(2):117–27.

2. Jaywant A, Toglia J, Gunning FM, O’Dell MW. Subgroups defined by the Montreal Cognitive Assessment differ in functional gain during acute inpatient stroke rehabilitation. Arch Phys Med Rehabil. 2019;

3. O’Dell MW, Jaywant A, Frantz M, Patel R, Kwong E, Wen K, et al. Changes in the Activity Measure for Post-Acute Care Domains in Persons With Stroke During the First Year After Discharge From Inpatient Rehabilitation. Arch Phys Med Rehabil [Internet]. 2021; Available from: https://doi.org/10.1016/j.apmr.2020.11.020

4. Jaywant A, Toglia J, Gunning FM, O’Dell MW. The clinical utility of a 30-minute neuropsychological assessment battery in inpatient stroke rehabilitation. J Neurol Sci [Internet]. 2018;390(April):54–62. Available from: https://doi.org/10.1016/j.jns.2018.04.012

5. Cahn-Weiner DA, Boyle PA, Malloy PF. Tests of executive function predict instrumental activities of daily living in community-dwelling older individuals. Appl Neuropsychol [Internet]. 2002 Jan;9(3):187–91. Available from: http://www.ncbi.nlm.nih.gov/pubmed/12584085

6. Toglia J. The Dynamic Interactional Model and the Multicontext Approach. In: Katz N, Toglia J, editors. Cogn Occup Across Lifesp Model Interv Occup Ther. Baltimore MD: American Occupational Therapy Association; 2018. p. 355–85.

7. Jaywant A, Steinberg C, Lee A, Toglia J. Feasibility and acceptability of the multicontext approach for individuals with acquired brain injury in acute inpatient rehabilitation: A single case series. Neuropsychol Rehabil [Internet]. 2020;1–20. Available from: https://doi.org/10.1080/09602011.2020.1810710

8. Toglia J, Askin G, Gerber LM, Taub MC, Mastrogiovanni AR, O’Dell MW. Association between 2 measures of cognitive instrumental activities of daily living and their relation to the Montreal Cognitive Assessment in persons with stroke. Arch Phys Med Rehabil [Internet]. 2017;98(11):2280–7. Available from: https://doi.org/10.1016/j.apmr.2017.04.007

9. Baum CM, Connor LT, Morrison T, Hahn M, Dromerick AW, Edwards DF. Reliability, validity, and clinical utility of the Executive Function Performance Test: A measure of executive function in a sample of people With stroke. Am J Occup Ther. 2008;62(4):446– 55.

10. Toglia J. Weekly Calendar Planning Activity (WCPA): A performance test of executive function. AOTA Press; 2015.

11. Toglia J, Lahav O, Ari E Ben, Kizony R. Adult age and cultural differences in performance on the Weekly Calendar Planning Activity (WCPA). Am J Occup Ther. 2017;71(5):1–7.

12. Goverover Y, Toglia J, DeLuca J. The weekly calendar planning activity in multiple sclerosis: A top-down assessment of executive functions. Neuropsychol Rehabil [Internet]. 2020;30(7):1372–87. Available from: https://doi.org/10.1080/09602011.2019.1584573

13. Lahav O, Katz N. Independent Older Adult’s IADL and Executive Function According to Cognitive Performance. OTJR Occup Particip Heal [Internet]. 2020;40(3):183–9. Available from: https://doi.org/10.1177/1539449220905813

14. Lahav O, Ben-Simon A, Inbar-Weiss N, Katz N. Weekly Calendar Planning Activity for University Students: Comparison of Individuals With and Without ADHD by Gender. J Atten Disord. 2018;22(4):368–78.

15. Doherty M. Validation of the Weekly Calendar Planning Activity with teenagers with acquired brain injury. Am J Occup Ther. 2018;72(4_Supplement_1):7211500026p1–7211500026p1.

16. Zlotnik S, Schiff A, Ravid S, Shahar E, Toglia J. A new approach for assessing executive functions in everyday life, among adolescents with Genetic Generalised Epilepsies. Neuropsychol Rehabil. 2018;30(2):333–45.

17. Kaizerman-Dinerman A, Roe D, Josman N. An efficacy study of a metacognitive group intervention for people with schizophrenia. Psychiatry Res [Internet]. 2018;270:1150–6. Available from: https://doi.org/10.1016/j.psychres.2018.10.037

18. Planton M, Peiffer S, Albucher JF, Barbeau EJ, Tardy J, Pastor J, et al. Neuropsychological outcome after a first symptomatic ischaemic stroke with “good recovery.” Eur J Neurol. 2012;19(2):212–9.

19. Zinn S, Bosworth HB, Hoenig HM, Swartzwelder HS. Executive function deficits in acute stroke. Arch Phys Med Rehabil. 2007;88(2):173–80.

20. Lussier A, Doherty M, Toglia J. Weekly Calendar Planning Activity. In: Functional Cognition and Occupational Therapy. AOTA Press; 2019. p. 75–91.

21. Nasreddine Z, Phillips N, Bédirian V, Charbonneau S, Whitehead V, Colllin I, et al. The Montreal Cognitive Assessment, MoCA: a brief screening tool for mild cognitive impairment. J Am Geriatr Soc [Internet]. 2005;53(4):695–9. Available from: http://onlinelibrary.wiley.com/doi/10.1111/j.1532-5415.2005.53221.x/full

22. Jaywant A, Toglia J, Gunning FM, O’Dell MW. The diagnostic accuracy of the Montreal Cognitive Assessment in inpatient stroke rehabilitation. Neuropsychol Rehabil. 2017;

23. Skidmore ER, Dawson DR, Butters MA, Grattan ES, Juengst SB, Whyte EM, et al. Strategy Training Shows Promise for Addressing Disability in the First 6 Months After Stroke. Neurorehabil Neural Repair. 2015;29(7):668–76.

24. Schiehser DM, Delis DC, Filoteo JV, Delano-Wood L, Han SD, Jak AJ, et al. Are self-reported symptoms of executive dysfunction associated with objective executive function performance following mild to moderate traumatic brain injury? J Clin Exp Neuropsychol. 2011;33(6):704–14.

